# Vaccination status and associated factors among healthcare workers providing primary care: a cross-sectional study in Yaoundé, Cameroon

**DOI:** 10.64898/2026.07.13.26357972

**Authors:** Ariane Nouko, Fabrice Zobel Lekeumo Cheuyem, Félicitée Nguefack, Chabeja Achangwa, Innocent Takougang

**Affiliations:** Department of Public Health, Faculty of Medicine and Biomedical Sciences, The University of Yaoundé 1, Yaoundé, Cameroon; Regional Delegation of Public Health, West Region, Bafoussam, Cameroon; Department of Pediatrics, Faculty of Medicine and Biomedical Sciences, The University of Yaoundé 1, Yaoundé, Cameroon; School of Biosciences, University of Kent, Canterbury, UK

**Keywords:** Healthcare workers, primary care, vaccination coverage, hepatitis B, tuberculosis, tetanus, occupational health, Cameroon

## Abstract

**Background:** Healthcare workers (HCWs) are at increased risk of exposure to vaccine-preventable diseases, including hepatitis B, tuberculosis (TB), and tetanus, due to their occupational environment. Despite the availability of safe and effective vaccines, vaccination coverage among HCWs remains suboptimal in many low- and middle-income countries. This study aimed to determine the vaccination coverage for hepatitis B, tuberculosis, and tetanus among HCWs in district hospitals in Yaoundé, Cameroon, and to identify factors associated with incomplete vaccination.

**Methods:** A cross-sectional analytical study was conducted from January to June 2024 across all seven district hospitals in Yaoundé. A total of 406 HCWs were enrolled using a structured, self-administered questionnaire. Incomplete vaccination status was defined as nit having received three doses for hepatitis B, at least two doses for TB, and at least three doses for tetanus. Data were analyzed using R Statistics version 4.3.3. Bivariate and multivariate logistic regression models were used to identify predictors of incomplete vaccination, with statistical significance set at *p* < 0.05.

**Results:** Most of participants were female (75.6%) and aged 30-44 years (65.5%). Full vaccination coverage was 36.2% for hepatitis B, 15.8% for tuberculosis, and 53.4% for tetanus. Unvaccinated proportions were 43.1% for hepatitis B, 0.5% for tuberculosis, and 17.2% for tetanus. Multivariate analysis revealed that HCWs aged 55-66 years were significantly less likely to be incompletely vaccinated against hepatitis B compared to those aged 19-29 years (aOR = 0.07; 95% CI: 0.01-0.39; *p* = 0.004). Nurses and laboratory technicians were more likely to be incompletely vaccinated than doctors (aOR = 7.06; 95% CI: 3.82-13.6; *p* < 0.001 and aOR = 3.78; 95% CI: 1.73-8.52; *p* = 0.001, respectively). Longer professional experience was protective against incomplete vaccination for all three vaccines (*p* < 0.05). The main barriers to vaccination included high cost, doubts about the need for vaccination, and concerns about side effects.

**Conclusions:** Vaccination coverage for hepatitis B, tuberculosis, and tetanus among HCWs in Yaoundé remains suboptimal, particularly for hepatitis B. Addressing financial barriers, improving access to vaccines, and implementing targeted educational interventions are essential to enhance vaccine uptake and protect both HCWs and their patients from preventable occupational infections.

## Introduction

Vaccination is one of the best and most affordable ways to prevent infectious diseases and lower illness and death rates around the world. In recent decades, global immunization programs have greatly reduced the incidence of many vaccine-preventable diseases and saved millions of lives each year [1,2]. Still, infectious diseases are a major public health problem, especially in low- and middle-income countries where healthcare resources are limited [1].

Healthcare workers (HCWs) play a central role in the prevention, diagnosis, and management of infectious diseases. However, the nature of their professional activities exposes them to numerous occupational hazards, including exposure to infectious agents through contact with patients, blood, and other biological fluids [3,4]. Needle-stick injuries, cuts from contaminated sharps, and exposure of mucous membranes to infected biological materials represent common routes of occupational transmission of blood-borne infections among healthcare personnel [3,5,6]. Globally, occupational exposure to contaminated sharps has been identified as a major contributor to infections such as hepatitis B virus (HBV), hepatitis C virus, and human immunodeficiency virus among healthcare workers [7]. These occupational hazards can also lead to tuberculosis (TB), resulting from exposure to droplets of expectoration from active contaminated patients, and to tetanus, due to lesions contaminated with tetanus spores, which are universally present in soil and on surfaces [8,9].

Hepatitis B is a major occupational hazard for healthcare workers worldwide, transmitted through contact with infected blood or fluids [3,5]. In 2022, about 254 million people had chronic HBV, causing over 1.1 million deaths annually from liver cirrhosis and cancer [10]. Healthcare workers face a high risk of this life-threatening disease due to exposure to contaminated materials [5,6,11]. Vaccination is highly effective and offers long-term protection, making it the key preventive measure against occupational HBV infection [12].

Tuberculosis (TB) poses a major global health risk and is an occupational hazard for healthcare workers, especially in high-burden countries. Healthcare workers in clinical settings frequently encounter active TB, raising exposure risks to *Mycobacterium tuberculosis*. Even with routine BCG vaccination during infancy in many countries, healthcare workers remain at risk, underscoring the need for infection prevention and control in healthcare facilities [13].

Tetanus is a vaccine-preventable disease caused by Clostridium tetani’s neurotoxin. Although not transmitted person-to-person, infection can occur from contaminated wounds. Healthcare workers risk exposure via injuries or contact with contaminated materials. Regular immunization with primary and booster doses is recommended for adults, including healthcare professionals, for occupational health [14].

Despite the availability of safe vaccines, vaccination rates among healthcare workers remain low in many countries. Studies show low vaccine uptake, especially in low-resource settings [15–17]. Barriers include limited access, lack of policies, insufficient knowledge, safety concerns, and costs. Factors like age, profession, and experience also affect vaccination behavior [6,15].

In sub-Saharan Africa, healthcare workers face high risks of infectious diseases due to common diseases and limited safety measures. Many in the region are unvaccinated against hepatitis B despite higher infection risks [18]. In Cameroon, several studies have reported low hepatitis B vaccination coverage among healthcare workers, with full vaccination rates ranging from approximately 13% to 27%, despite high occupational risk [5,6,19,20]. In addition, occupational exposure to blood and body fluids among healthcare workers is frequent, further increasing the risk of infection [11,21,22].

While hepatitis B vaccination among healthcare workers has been relatively well studied, there is limited evidence regarding tuberculosis and tetanus vaccination coverage among healthcare workers in Cameroon and similar low-resource settings. This highlights an important gap in the literature and underscores the need for a comprehensive assessment of vaccination status across multiple vaccine-preventable diseases.

Ensuring healthcare workers are properly vaccinated is crucial for protecting both the staff and their patients. Vaccination reduces the risk of healthcare workers contracting infections at work and helps prevent the spread of diseases in healthcare settings [4]. However, there is still limited information on how many healthcare workers in Cameroon are vaccinated or what barriers that prevent them to get vaccinated. This study aimed to determine how many healthcare workers in district hospitals in Yaoundé are vaccinated against hepatitis B, tuberculosis, and tetanus, and to identify factors associated with incomplete vaccination.

## Methods

### Study design and period

We conducted a hospital-based cross-sectional and analytical study in the seven (7) DHs in Yaoundé from January to June 2024.

### Study setting

Yaoundé serves as the political capital of Cameroon and the Centre region. Its population was projected to be 4.85 million by 2025. It is the nation’s second-biggest city, and all ethnic groups in Cameroon are effectively represented there. The health system in Cameroon is structured around health districts, serving as the operational level for delivering healthcare. A district hospital serves as the initial level of reference in the healthcare hierarchy and delivers primary medical care. The seven DHs in Yaoundé, which include Biyem-Assi, Cité-Verte, Djoungolo-Olembe, Efoulan, Mvog-Ada, Nkolndongo, and Odza, serve around 4.2 million people, employing almost 560 health workers, offering 650 beds, and managing 216,235 consultations and 25,320 admissions each year

### Participants

Our study population included all healthcare workers providing care to the general population in Yaoundé health facilities. Meanwhile, study participants included HCWs from the following clinical units at each DH: surgery, internal medicine, obstetrics and gynecology, laboratory, pediatrics, emergency, outpatient department, and hygiene department who provided written informed consent to participate in this study.

### Sampling method

The sample size was calculated using the single proportion formula (n=[Z_α/2_]^2^ *[P (1-P)]/E^2^) at a 95% level of confidence, where Z_α/2_=1.96 and P=34.0% [5]. Using a margin error of E=5%, the minimum sample size obtained was: 345. Adding a 10% dropout rate, the final minimum sample size was: 384 participants. In each hospital department, all consenting personnel were enrolled in the study.

### Data collection

An anonymous, structured, self-administered questionnaire consisting of 25 questions organized into two main sections was developed and used to collect data. It captured information on socio-demographic characteristics, vaccination coverage, reasons for non-vaccination, and major side effects of each vaccine (**Supplementary Material: Questionnaire**).

### Variable and operational definition

Independent variables included sex, age group, professional experience in years, health unit in which HCWs worked, and the HCWs’ grade. The dependent variable was vaccination coverage for Hepatitis B, Tuberculosis, and tetanus vaccines. HCWs were considered fully vaccinated if they had completed 3 doses of the Hepatitis B vaccine, at least 2 doses of the tuberculosis vaccine, and at least 3 doses of the tetanus vaccine.

### Data processing and analysis

Every completed questionnaire was verified, inputted, recoded as required, and analyzed utilizing R Statistics Version 4.3.3. Categorical variables were characterized by frequency (n) and percentage (%). The Fisher’s exact test was employed to compare ratios. Simple and multiple binary logistic regressions were employed to evaluate the strength of the association between variables and to adjust for possible confounding factors. The predictors that best fit the model were selected gradually using the Akaike Information Criterion (AIC). The model having the smallest index was chosen. A *p*-value < 0.05 was considered statistically significant. Confidence Intervals (CIs) were calculated at a 95% confidence level.

### Ethical considerations

The Institutional Review Board of the Regional Delegation of Public Health for the Center Region (No 0244-3/CRERSHC/2024) and the Faculty of Medicine and Biomedical Sciences IRB (No 1115/UY1/FMSB/VDRC/DAASR/CSD) authorized this investigation. Before being included in the study, each subject gave their informed consent. Every procedure was carried out in compliance with the Declaration of Helsinki.

## Results

### Socio-professional characteristics of study participants

Out of the 450 HCWs contacted, 435 gave their consent, 406 returned the completed questionnaire, representing a 90% response rate. Most of our study participants were female (75.6%). Participants aged 30-45 years were the most represented (65.5%). They were mostly nurses (61.3%) and medical doctors (17.0%) (**Table 1**).

**Table 1.** Socio-professional characteristics of study participants, Yaoundé District Hospitals, June 2024 (n = 406)

| Characteristic | Count (n) | Frequency (%) |
| --- | --- | --- |
| **Age group (in years)** |  |  |
| 19-29 | 55 | 13.5 |
| 30-44 | 266 | 65.5 |
| 45-54 | 74 | 18.2 |
| 55-66 | 11 | 2.7 |
| **Sex** |  |  |
| Female | 307 | 75.6 |
| Male | 99 | 24.4 |
| **Level of education** |  |  |
| Primary | 3 | 0.7 |
| Secondary | 84 | 20.7 |
| Tertiary | 319 | 78.6 |
| **Professional grade** |  |  |
| Medical student | 4 | 1.0 |
| Nurse | 249 | 61.3 |
| Doctor | 69 | 17.0 |
| Administrative staff | 4 | 1.0 |
| Hygiene staff | 14 | 3.4 |
| Laboratory technician | 54 | 13.3 |
| Other technicians | 12 | 3.0 |
| **Professional experience (in years)** |  |  |
| 1-4 | 116 | 28.6 |
| 5-14 | 224 | 55.2 |
| 15-34 | 66 | 16.3 |
| **Department** |  |  |
| Dentistry | 18 | 4.4 |
| Laboratory | 48 | 11.8 |
| Internal medicine | 63 | 15.5 |
| Paediatrics | 40 | 9.9 |
| Maternity and gynaecology | 54 | 13.3 |
| Surgery | 46 | 9.9 |
| Outpatient (vaccination) | 30 | 7.4 |
| Hygiene | 15 | 3.7 |
| Others (OPD consultation, emergency, etc) | 92 | 22.7 |

**OPD: Outpatient Department**

### Vaccination coverage

Close to half (43.1%) of the HCWs in Yaoundé had not taken viral hepatitis B vaccine, while just 36.2% were fully vaccinated against viral hepatitis B vaccine. Most of the HCWs (83.7%) had taken just one dose of the tuberculosis vaccine at birth, and just 15.8% had taken more than one dose. Half of the HCWs (53.4%) reported to have completed their tetanus vaccine, while 29.3% had taken incomplete doses. The compliance with vaccination among HCWs across various District Hospital of Yaoundé was almost homogenous as no statistically significant differences was observed between different groups (**Table 2**).

**Table 2.** Viral hepatitis B, tuberculosis, and tetanus vaccination coverage among healthcare workers in Yaoundé District Hospitals, June 2024.

| Type of vaccine | District hospital | Fully | Partially n (%) | Unvaccinated | Total (100%) | p-value<sup>1</sup> |
| --- | --- | --- | --- | --- | --- | --- |
| Hepatitis B |  |  |  |  |  | 0.327 |
|  | Biyem-Assi | 27 (40.9) | 12 (18.2) | 27 (40.9) | 66 |  |
|  | Cite-Verte | 16 (26.2) | 17 (27.9) | 28 (45.9) | 61 |  |
|  | Djoungolo | 21 (35.0) | 14 (23.3) | 25 (41.7) | 60 |  |
|  | Efoulan | 30 (46.2) | 8 (12.3) | 27 (41.5) | 65 |  |
|  | Mvog-Ada | 17 (32.7) | 12 (23.1) | 23 (44.2) | 52 |  |
|  | Nkolndongo | 13 (25.0) | 13 (25.0) | 26 (50.0) | 52 |  |
|  | Odza | 23 (46.0) | 8 (16.0) | 19 (38.0) | 50 |  |
| **Total** |  | 147 (36.2) | 84 (20.7) | 175 (43.1) | 406 |  |
| Tuberculosis |  |  |  |  |  | 0.125 |
|  | Biyem-Assi | 13 (19.7) | 53 (80.3) | 0 (0.0) | 66 |  |
|  | Cite-Verte | 10 (16.4) | 51 (83.6) | 0 (0.0) | 61 |  |
|  | Djoungolo | 8 (13.3) | 51 (85.0) | 1 (1.7) | 60 |  |
|  | Efoulan | 8 (12.3) | 57 (87.7) | 0 (0.0) | 65 |  |
|  | Mvog-Ada | 4 (7.7) | 48 (92.3) | 0 (0.0) | 52 |  |
|  | Nkolndongo | 6 (11.5) | 45 (86.5) | 1 (1.9) | 52 |  |
|  | Odza | 15 (30.0) | 35 (70.0) | 0 (0.0) | 50 |  |
| **Total** |  | 64 (15.8) | 340 (83.7) | 2 (0.5) | 406 |  |
| Tetanus |  |  |  |  |  | 0.224 |
|  | Biyem-Assi | 41 (62.1) | 14 (21.2) | 11 (16.7) | 66 |  |
|  | Cite-Verte | 23 (37.7) | 22 (36.1) | 16 (26.2) | 61 |  |
|  | Djoungolo | 29 (48.3) | 19 (31.7) | 12 (20.0) | 60 |  |
|  | Efoulan | 37 (56.9) | 20 (30.8) | 8 (12.3) | 65 |  |
|  | Mvog-Ada | 32 (61.5) | 10 (19.2) | 10 (19.2) | 52 |  |
|  | Nkolndongo | 27 (51.9) | 18 (34.6) | 7 (13.5) | 52 |  |
|  | Odza | 28 (56.0) | 16 (32.0) | 6 (12.0) | 50 |  |
| **Total** |  | 217 (53.4) | 119 (29.3) | 70 (17.2) | 406 |  |

1: Fisher exact probability test

### Predictors of complete vaccination

Multivariate analysis identified age, professional experience and professional grade as best predictor of viral hepatitis B complete vaccination.

Participants aged 55-66 were significantly fourteen times less likely to be unvaccinated against the viral hepatitis B compared to those aged 19-30 (aOR = 0.07; 95%CI: 0.01-0.39; *p* = 0.004). Similarly, HCW with more professional (5-14 years) experience were significantly twice less likely to be non-fully vaccinated against viral hepatitis B (aOR = 0.51; 95%CI: 0.25-0.97; *p* = 0.044) compared to HCW with lower professional experience. Conversely, nurses and laboratory technician were significantly seven times (aOR = 7.06; 95% CI: 3.82-13.6; *p* < 0.001) and four times (aOR = 3.78; 95% CI: 1.73-8.52; *p* = 0.001) more likely to be non-fully vaccinated against the viral hepatitis B compared to doctors respectively (**Table 3**).

**Table 3.** Simple and multiple binary logistic regressions of factors associated with incomplete vaccination against viral hepatitis B among healthcare workers in Yaoundé District Hospitals, June 2024 (n = 406)

| Predictor | Incomplete vaccination |  | *p*-value<sup>1</sup> | cO | aO | 95% CI limits |  | *p*-value |
| --- | --- | --- | --- | --- | --- | --- | --- | --- |
|  | n (%) |  |  | R | R |  |  |  |
|  | Yes | No |  |  |  | Lower | Upper |  |
| Age group (in years) |  |  | 0.009 |  |  |  |  |  |
| 19-30 | 12 (21.8) | 43 (78.2) |  | 1 |  |  |  |  |
| 30-44 | 98 (36.8) | 168 (63.2) |  | 0.48 | 0.48 | 0.19 | 1.20 | 0.122 |
| 45-54 | 29 (39.2) | 45 (60.8) |  | 0.43 | 0.36 | 0.11 | 1.14 | 0.086 |
| 55-66 | 8 (72.7) | 3 (27.3) |  | 0.10 | 0.07 | 0.01 | 0.39 | 0.004 |
| **Sex** |  |  | 0.472 |  |  |  |  |  |
| Male | 39 (39.4) | 60 (60.6) |  | 1 |  |  |  |  |
| Female | 108 (35.2) | 199 (64.8) |  | 1.20 |  |  |  |  |
| **Professional experience (in years)** |  |  | 0.011 |  |  |  |  |  |
| 1-4 | 29 (25.0) | 87 (75.0) |  | 1 | 1 |  |  |  |
| 5-14 | 91 (40.6) | 133 (59.4) |  | 0.49 | 0.51 | 0.25 | 0.97 | 0.044 |
| 15-34 | 27 (40.9) | 39 (59.1) |  | 0.48 | 0.72 | 0.27 | 1.93 | 0.517 |
| **Department** |  |  | 0.292 |  |  |  |  |  |
| Outpatient (vaccination) | 15 (50.0) | 15 (50.0) |  | 1 |  |  |  |  |
| Dentistry | 7 (38.9) | 11 (61.1) |  | 1.57 |  |  |  |  |
| Hygiene | 1 (6.7) | 14 (93.3) |  | 14.0 |  |  |  |  |
| Laboratory | 21 (43.8) | 27 (56.3) |  | 1.29 |  |  |  |  |
| Internal medicine | 29 (46.0) | 34 (54.0) |  | 1.17 |  |  |  |  |
| Gynecology | 17 (31.5) | 37 (68.5) |  | 2.18 |  |  |  |  |
| Pediatrics | 14 (35.0) | 26 (65.0) |  | 1.86 |  |  |  |  |
| Surgery | 16 (34.8) | 30 (65.2) |  | 1.88 |  |  |  |  |
| Others units | 27 (29.3) | 65 (70.7) |  | 2.41 |  |  |  |  |
| **Professional grade** |  |  | < 0.001 |  |  |  |  |  |
| Doctor | 46 (66.7) | 23 (33.3) |  | 1 |  |  |  |  |
| Medical student | 1 (25.0) | 3 (75.0) |  | 6.00 | 2.95 | 0.33 | 63.3 | 0.371 |
| Laboratory technician | 24 (44.4) | 30 (55.6) |  | 2.50 | 3.78 | 1.73 | 8.52 | 0.001 |
| Nurse | 71 (28.5) | 178 (71.5) |  | 5.01 | 7.06 | 3.82 | 13.6 | <0.001 |
| Hygiene staff | 1 (7.1) | 13 (92.9) |  | 26.0 | 42.0 | 7.36 | 798 | <0.001 |
| Other staff | 1 (25.0) | 3 (75.0) |  | 6.00 | 7.43 | 0.83 | 160 | 0.099 |
| Other technician | 3 (25.0) | 9 (75.0) |  | 6.00 | 7.92 | 2.04 | 39.6 | 0.005 |

1: Fisher exact test; cOR: Crude odds ratio; aOR = Adjusted odds ratio; CI = Confidence interval

The multivariate analysis identified the professional experience as the best unique predictor of non-complete vaccination against tuberculosis. HCWs with more experience were significantly four times (aOR = 0.28; 95%CI: 0.11-0.65; *p* = 0.006) and nine times (aOR = 0.11; 95%CI: 0.04-0.25; *p* < 0.001) respectively less likely to be incompletely vaccinated against tuberculosis (**Table 4**).

**Table 4.**
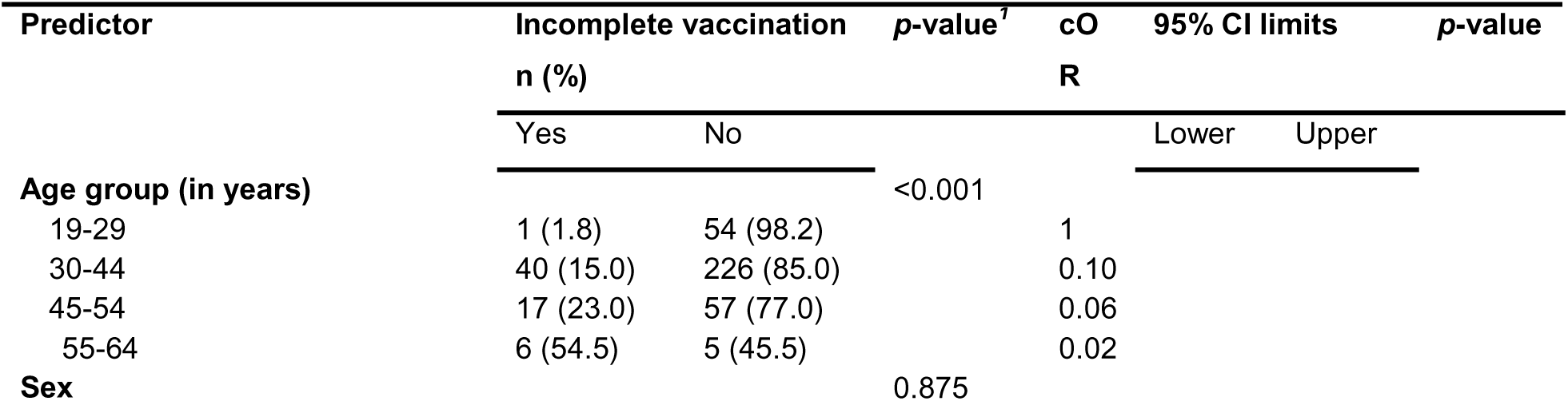
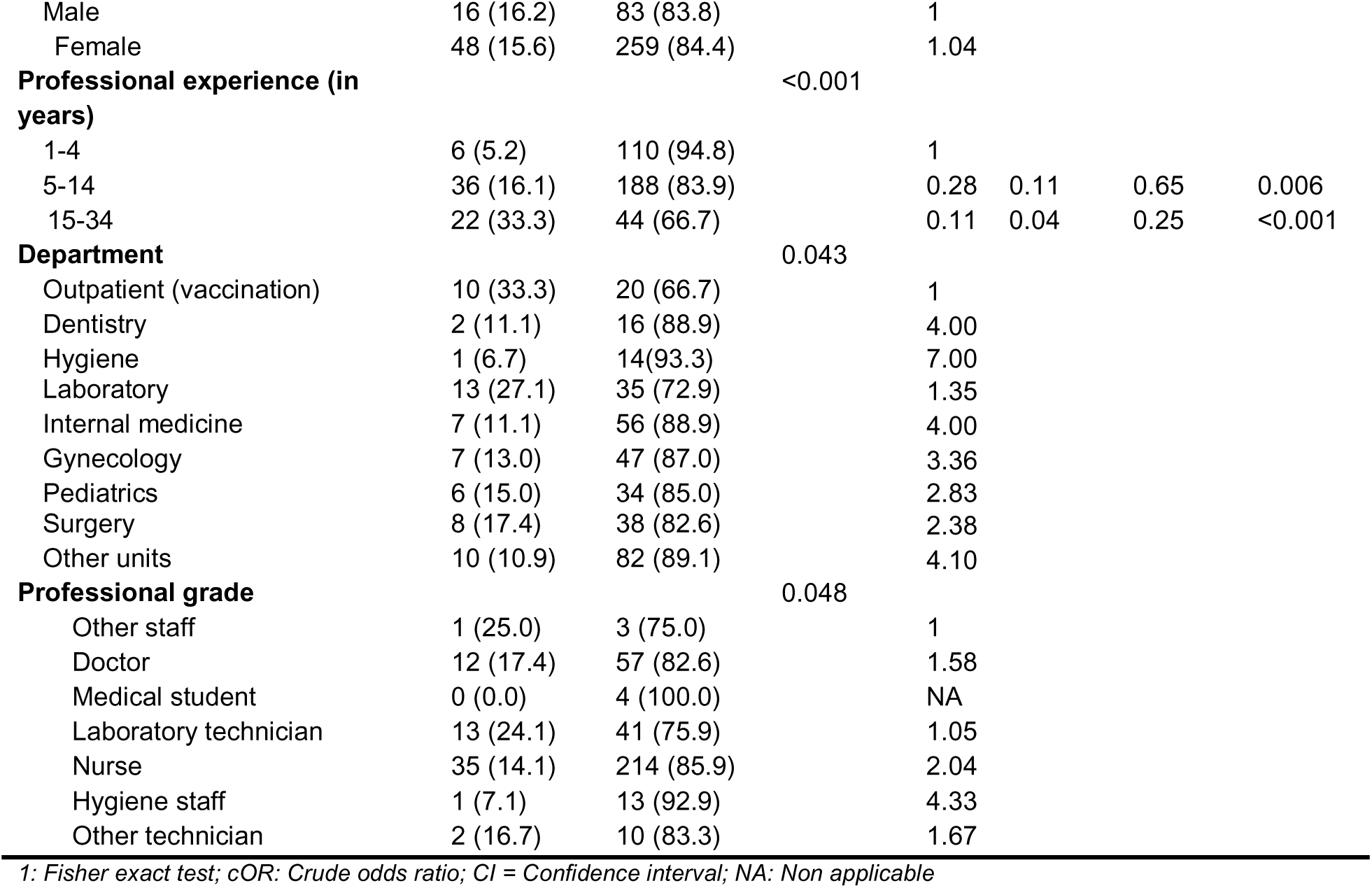
Simple and multiple binary logistic regressions of factors associated with incomplete vaccination against tuberculosis among healthcare workers in Yaoundé District Hospitals, June 2024 (n = 406)

The multivariate analysis revealed that older age was significantly associated with 3-16 time less risk of being non-fully immunized against tetanus (*p* < 0,05) compared to younger HCWs (19-29 years). Although non-significant, male HCWs were 66% more likely to be non-fully vaccinated against tetanus (aOR = 1.66; 95%CI: 0.00-0.56; *p* = 0.055). HCWs with more years of professional experience were significantly less likely (6-9 times) to be incompletely vaccinated against tetanus (*p* < 0,05) compared to those with less years of professional experience (1-4 years) (**Table 5**).

**Table 5.** Simple and multiple binary logistic regressions of factors associated with incomplete vaccination against tetanus among healthcare workers in Yaoundé District Hospitals, June 2024 (n = 406)

| Predictor | Incomplete vaccination |  | *p*-value<sup>1</sup> | cO | aO | 95% CI limits |  | *p*-value |
| --- | --- | --- | --- | --- | --- | --- | --- | --- |
|  | n (%) |  |  | R | R |  |  |  |
|  | Yes | No |  |  |  | Lower | Upper |  |
| Age group (in years) |  |  | <0.001 |  |  |  |  |  |
| 19-29 | 5 (9.1) | 50 (90.9) |  | 1 | 1 |  |  |  |
| 30-44 | 140 (52.6) | 126 (47.4) |  | 0.09 | 0.30 | 0.09 | 0.82 | 0.027 |
| 45-54 | 62 (83.8) | 12 (16.2) |  | 0.02 | 0.10 | 0.03 | 0.37 | <0.001 |
| 55-56 | 10 (90.9) | 1 (9.1) |  | 0.01 | 0.06 | 0.00 | 0.56 | 0.028 |
| **Sex** |  |  | 0.297 |  |  |  |  |  |
| Female | 48 (48.5) | 51 (51.5) |  | 1 | 1 |  |  |  |
| Male | 169 (55.0) | 138 (45.0) |  | 1.30 | 1.66 | 0.99 | 2.80 | 0.055 |
| **Professional experience (in years)** |  |  | <0.001 |  |  |  |  |  |
| 1-4 | 20 (17.2) | 96 (82.8) |  | 1 | 1 |  |  |  |
| 5-14 | 143 (63.8) | 81 (36.2) |  | 0.12 | 0.19 | 0.10 | 0.36 | <0.001 |
| 15-34 | 54 (81.8) | 12 (18.2) |  | 0.05 | 0.16 | 0.06 | 0.43 | <0.001 |
| **Department** |  |  | 0.325 |  |  |  |  |  |
| Outpatient (vaccination) | 20 (66.7) | 10 (33.3) |  | 1 |  |  |  |  |
| Dentistry | 7 (38.9) | 11 (61.1) |  | 3.14 |  |  |  |  |
| hygiene | 4 (26.7) | 11 (73.3) |  | 5.50 |  |  |  |  |
| Laboratory | 28 (58.3) | 20 (41.7) |  | 1.43 |  |  |  |  |
| Medicine | 32 (50.8) | 31 (49.2) |  | 1.94 |  |  |  |  |
| Gynecology | 28 (51.9) | 26 (48.1) |  | 1.86 |  |  |  |  |
| Pediatrics | 21 (52.5) | 19 (47.5) |  | 1.81 |  |  |  |  |
| Surgery | 27 (58.7) | 19 (41.3) |  | 1.41 |  |  |  |  |
| Others | 50 (54.3) | 42 (45.7) |  | 1.68 |  |  |  |  |
| **Professional grade** |  |  | 0,161 |  |  |  |  |  |
| Laboratory technician | 32 (59.3) | 22 (40.7) |  | 1 |  |  |  |  |
| Doctor | 36 (52.2) | 33 (47.8) |  | 1.33 |  |  |  |  |
| Medical student | 0 (0.0) | 4 (100.0) |  | NA |  |  |  |  |
| Nurse | 137 (55.0) | 112 (45.0) |  | 1.19 |  |  |  |  |
| Hygiene staff | 4 (28.6) | 10 (71.4) |  | 3.64 |  |  |  |  |
| Other staff | 2 (50.0) | 2 (50.0) |  | 1.45 |  |  |  |  |
| Other technician | 6 (50.0) | 6 (50.0) |  | 1.45 |  |  |  |  |

1: Fisher exact test; cOR: Crude odds ratio; aOR = Adjusted odds ratio; CI = Confidence interval; NA: Non applicable

### Main reported adverse effects

HCWs reported fever, headache and large sore as the main adverse effects observed after having received the vaccines against the viral hepatitis B, tuberculosis, and tetanus vaccines (**Fig. 1**).

**Fig. 1.**
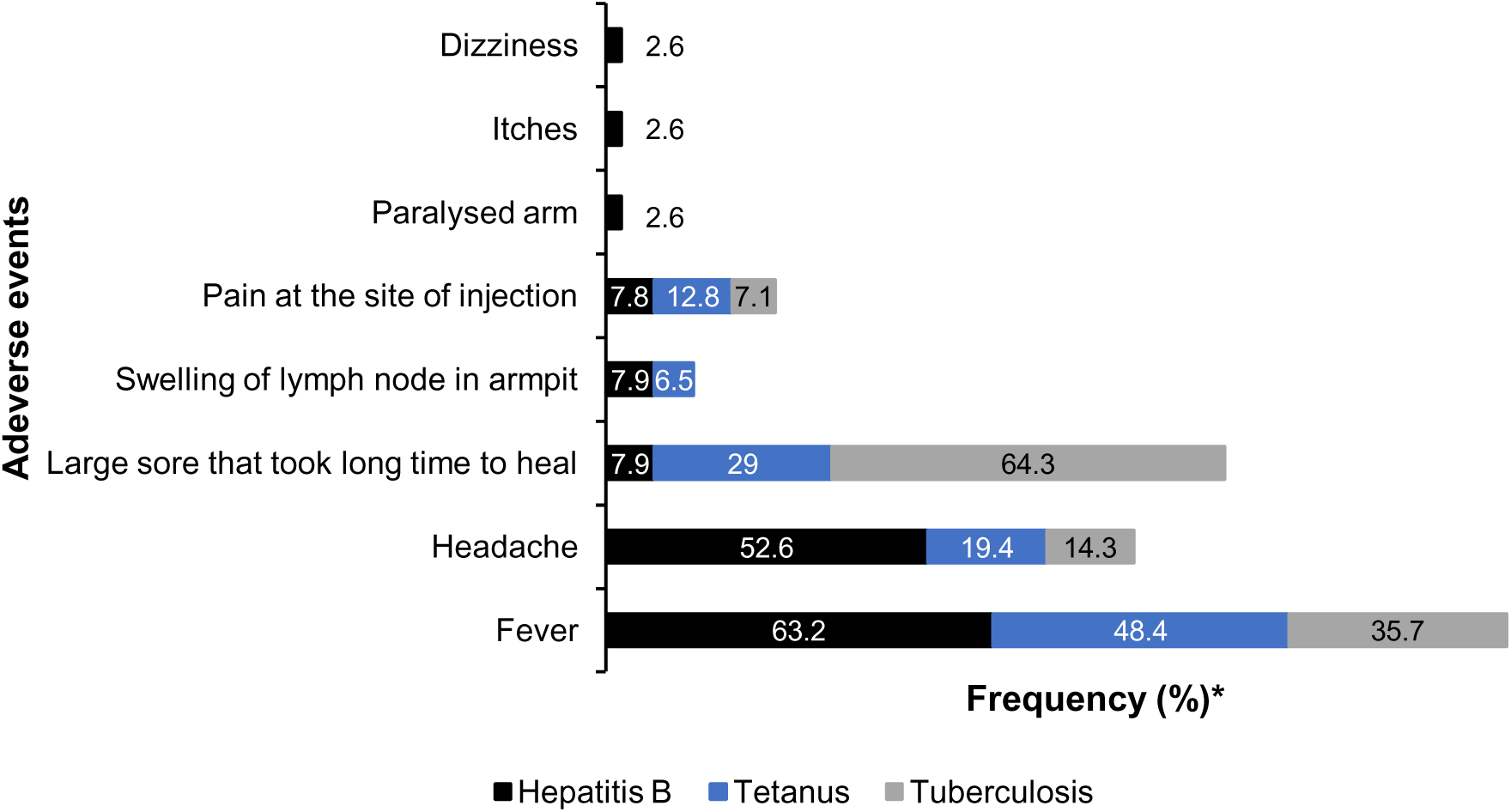
Main reported adverse effects per vaccine among healthcare workers in Yaoundé District Hospital, June 2024 (* For each specific vaccine received; n = 231 for viral hepatitis B vaccine; n = 404 for tetanus vaccine, and n = 336 for tuberculosis vaccine)

### Main reasons for non-vaccination

The main reasons for non-vaccinations for the Hepatitis B, TB, and Tetanus vaccines were high cost, doubts about the need for the vaccine and vaccine’s side effects (**Fig. 2**).

**Fig. 2.**
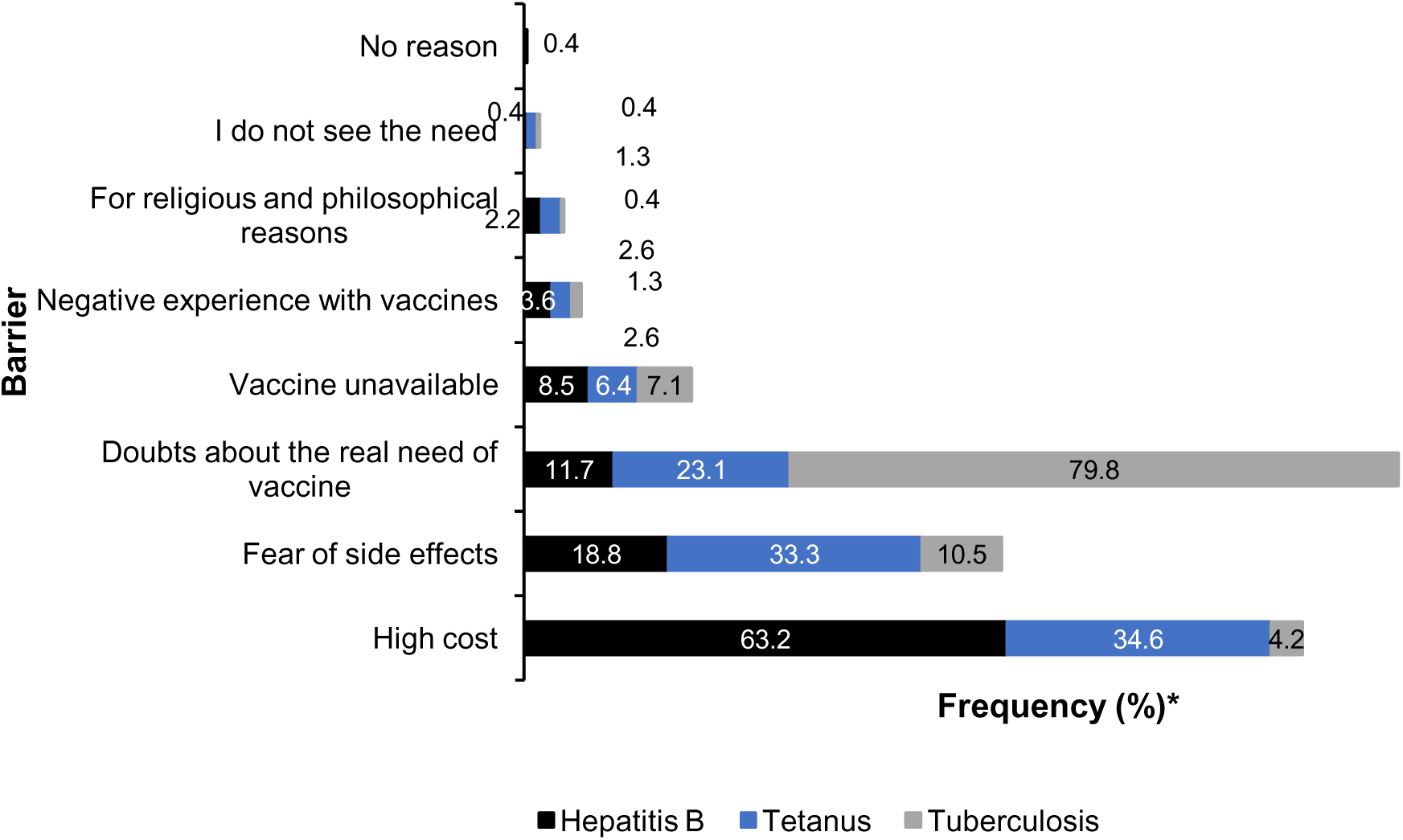
Main reported barriers for non-vaccination per vaccine among healthcare workers in Yaoundé District Hospital, June 2024 *(* For each specific vaccine not received; n = 231 for viral hepatitis B vaccine; n = 404 for tetanus vaccine, and n = 336 for tuberculosis vaccine)*

## Discussion

This study examined the proportion of healthcare workers at district hospitals in Yaoundé who were vaccinated against hepatitis B, tuberculosis, and tetanus, and the factors contributing to incomplete vaccination. The results indicate that vaccination coverage remains low, especially for hepatitis B, despite the high risks healthcare workers face at work.

In this study, only 36.2% of healthcare workers were fully vaccinated against hepatitis B, while 43.1% had not been vaccinated at all. This aligns with other research from sub-Saharan Africa, where hepatitis B vaccination rates among healthcare workers are also low, with studies reporting as low as 24.7%, indicating that a large proportion of healthcare workers remain unprotected [17]. Previous Studies from African countries also report low vaccination rates, often below 50% [23]. Importantly, findings from Cameroon show even lower vaccination coverage among healthcare workers. A national study reported that fewer than one in six healthcare workers were fully vaccinated against hepatitis B, while about one in twelve had evidence of infection [20]. Other studies in Cameroon have also reported vaccination coverage as low as 4.7% and around 34%, confirming that inadequate vaccination remains a major concern in the country [5,11,19,24]. These findings reinforce the results of this study and highlight the persistence of low vaccination coverage at both national and regional levels.

The high proportion of unvaccinated healthcare workers in this study is major public health issues because they often face risk at work through blood and body fluid exposure. This low coverage represents a significant occupational health concern. In Cameroon, healthcare workers are often exposed to blood and other body fluids, increasing the risk of blood-borne infections. A recent review found that many healthcare workers in Cameroon have been exposed at work, underscoring the urgent need for preventive measures such as vaccination [21]. Also, most healthcare workers reported having received one dose of the tuberculosis vaccine, likely given at birth (83.7%). This reflects common practice in many low- and middle-income countries, where BCG vaccination is administered to infants. However, BCG does not provide strong protection for adults, especially against pulmonary tuberculosis. Therefore, even with high vaccination rates, healthcare workers remain at risk during work, highlighting the importance of other infection prevention and control measures to limit the spread of this infection.

In this study, more than half of the participants reported complete tetanus vaccination, although a considerable proportion remained partially vaccinated or unvaccinated. While tetanus vaccination coverage was higher than hepatitis B vaccination coverage, incomplete vaccination suggests gaps in adherence to booster-dose recommendations. This finding indicates the need for improved adult immunization strategies targeting healthcare workers, particularly regarding booster dose compliance.

In our study, age, work experience, and job type were associated with hepatitis B vaccination rates. Older and more experienced healthcare workers were more likely to be fully vaccinated. This may reflect increased awareness of occupational risks, greater exposure to vaccination campaigns, or accumulated experience within the healthcare system. Our findings are consistent with previous studies in Africa, which show that greater experience leads to higher vaccination rates due to greater awareness of risks [17,25]. On the other hand, nurses and lab technicians were less likely to be fully vaccinated than doctors. Other research has also found that non-physician healthcare workers often have lower vaccination rates, possibly because they have less access to information, support and insufficient financial resources as this vaccine as nearly two-third raised the high cost as the main barrier to vaccination against this disease especially for those borne before 2005 period of hepatitis B vaccine introduction in the routine immunization service in Cameroon [26].

Professional experience was also identified as a significant predictor of vaccination status for tuberculosis and tetanus. Healthcare workers with more years of experience were less likely to have incomplete vaccination, suggesting that experience plays a key role in shaping preventive health behaviors. These findings are consistent with studies conducted in low-resource settings, which have shown that more experienced healthcare workers are more likely to adopt protective measures, including vaccination [18].

The study identified several key barriers to vaccination against hepatitis B, tetanus, and tuberculosis. The main reasons for not getting vaccinated were the high cost of vaccines, doubts about the necessity of vaccination, and worries about side effects. These findings are consistent with other studies in sub-Saharan Africa, where cost, limited access, and lack of awareness are significant barriers for healthcare workers [15]. Finance remains a major challenge in low-resource settings, where healthcare workers often have to pay for vaccines out of pocket, thereby limiting access. In addition, misconceptions about vaccine safety and limited awareness of occupational risks may further contribute to low vaccination coverage.

HCWs reported mild side effects, such as fever, headache, and local pain, which are common with vaccines. However, concerns about these side effects may make some people hesitant to get vaccinated or to complete their vaccination series. Addressing these concerns through targeted education and awareness campaigns is essential to improving vaccine acceptance among healthcare personnel.

This study’s findings should be viewed considering Cameroon’s high occupational exposure to infectious risks among healthcare worker and the high prevalence of hepatitis B in Cameroon [21,27–29]. A recent systematic review shows that many healthcare workers face exposure to blood and body fluids, raising their risk of blood-borne infections [21]. Therefore, improving vaccination coverage among healthcare workers in Cameroon is crucial for occupational health and infection prevention.

## Limitations

This study has some limitations. Vaccination status was self-reported and may be subject to recall or social desirability bias. The cross-sectional design limits causal inference. In addition, the study was conducted in selected hospitals in Yaoundé, which may limit generalizability.

## Conclusions

Vaccination coverage among healthcare workers in Yaoundé remains suboptimal, particularly for hepatitis B. Factors such as age, professional experience, and job category influence vaccination status, while cost, limited access, and vaccine-related concerns remain key barriers. Strengthening vaccination programs through improved access, institutional policies, and awareness is essential to enhance the protection of healthcare workers.

## Abbreviations

AIC: Akaike Information Criterion
aOR: Adjusted odds ratio
BCG: Bacillus Calmette-Guérin (tuberculosis vaccine)
CI: Confidence interval
cOR: Crude odds ratio
DH: District Hospital
HBV: Hepatitis B virus
HCW: Healthcare worker
IRB: Institutional Review Board
NA: Not applicable
OPD: Outpatient Department
TB: Tuberculosis

## Data Availability

The datasets used and/or analyzed during the current study are available from the corresponding author on reasonable request.

## Acknowledgements

Our gratitude goes to all the healthcare workers who agreed to participate in this study and to the managers of the health facilities who gave their authorization for the conduct of this study.

## Author’s contribution

Drafting of the study protocol, data collection, analysis and interpretation, drafting and editing of manuscript: AN, FZLC, and AC; Critical revision of protocol, critical revision of manuscript: AN, FZLC, FN, and IT; Conception, design and supervision of research protocol and implementation, data analysis plan, revision, editing and final validation of the manuscript: IT.

## Declarations

### Consent for publication

Not applicable.

### Authors’ details

^1^ Department of Public Health, Faculty of Medicine and Biomedical Sciences, The University of Yaoundé 1, Yaoundé, Cameroon

^2^ Regional Delegation of Public Health, West Region, Bafoussam, Cameroon

^3^ Department of Pediatrics, Faculty of Medicine and Biomedical Sciences, The University of Yaoundé 1, Yaoundé, Cameroon

^4^ Department of Public Health, Faculty of Medical Sciences, University of the West Indies, Bridgetown, Barbados

## References

1. Immunization coverage. WHO, Geneva. 2025. https://www.who.int/news-room/fact-sheets/detail/immunization-coverage. Accessed 16 Mar 2026

2. Andre FE, Booy R, Bock HL, Clemens J, Datta SK, John TJ, et al. Vaccination greatly reduces disease, disability, death and inequity worldwide. Bull World Health Organ. 2008; 86:140–6. 10.2471/blt.07.040089

3. Beltrami EM, Williams IT, Shapiro CN, Chamberland ME. Risk and management of blood-borne infections in health care workers. Clin Microbiol Rev. 2000; 13:385–407. 10.1128/CMR.13.3.385

4. Haviari S, Bénet T, Saadatian-Elahi M, André P, Loulergue P, Vanhems P. Vaccination of healthcare workers: A review. Hum Vaccines Immunother. 2015; 11:2522–37. 10.1080/21645515.2015.1082014

5. Cheuyem FZL, Lyonga EE, Kamga HG, Mbopi-Keou F-X, Takougang I. Needlestick and Sharp Injuries and Hepatitis B Vaccination among Healthcare Workers: A Cross-Sectional Study in Six District Hospitals in Yaounde (Cameroon). J Community Med Public Health. Gavin Publishers; 2023; 7:1–9. 10.29011/2577-2228.100321

6. Cheuyem FZL, Lyonga EE, Takougang I. Standard precautions perception and practice among health workers in the obstetrics-gynecology department of a referral hospital in Cameroon. BMC Health Serv Res. 2025; 25:1165. 10.1186/s12913-025-13448-4

7. Prüss-Ustün A, Rapiti E, Hutin Y. Estimation of the global burden of disease attributable to contaminated sharps injuries among health-care workers. Am J Ind Med. 2005; 48:482–90. 10.1002/ajim.20230

8. Matuka DO, Duba T, Ngcobo Z, Made F, Muleba L, Nthoke T, et al. Occupational Risk of Airborne Mycobacterium tuberculosis Exposure: A Situational Analysis in a Three-Tier Public Healthcare System in South Africa. Int J Environ Res Public Health. 2021; 18:10130. 10.3390/ijerph181910130

9. Sudarshan R, Sayo AR, Renner DR, Saram S de, Godbole G, Warrell C, et al. Tetanus: recognition and management. Lancet Infect Dis. Elsevier; 2025;25: e645–57. 10.1016/S1473-3099(25)00292-0

10. Hiebert-Suwondo L, Manning J, Tohme RA, Buti M, Kondili LA, Spearman CW, et al. A 2024 global report on national policy, programmes, and progress towards hepatitis B elimination: findings from 33 hepatitis elimination profiles. Lancet Gastroenterol Hepatol. 2025; 10:671–84. 10.1016/S2468-1253(25)00069-X

11. Takougang I, Cheuyem FZL, Ze BRS, Tsamoh FF, Moneboulou HM. Awareness of standard precautions, circumstances of occurrence and management of occupational exposures to body fluids among healthcare workers in a regional level referral hospital (Bertoua, Cameroon). BMC Health Serv Res. 2024; 24:424. 10.1186/s12913-024-10855-x

12. Batra V, Goswami A, Dadhich S, Kothari D, Bhargava N. Hepatitis B immunization in healthcare workers. Ann Gastroenterol. 2015; 28:276–80.

13. Dye C. Global epidemiology of tuberculosis. Lancet. London, England; 2006;367:938–40. 10.1016/S0140-6736(06)68384-0

14. World Health Organization. Electronic address. Tetanus vaccines: WHO position paper, February 2017 - Recommendations. Vaccine. 2018; 36:3573–5. 10.1016/j.vaccine.2017.02.034

15. Kroflin K, Gonzalez Utrilla M, Moore M, Lomazzi M. Protecting the healthcare workers in low- and lower-middle-income countries through vaccination: barriers, leverages, and next steps. Glob Health Action. 2023; 16:2239031. 10.1080/16549716.2023.2239031

16. Gaviola GC, McCarville M, Shendale S, Goodman T, Lomazzi M, Desai S. A review of health worker vaccination programs in low, middle and upper middle-income countries. Public Health Pract. 2023; 6:100415. 10.1016/j.puhip.2023.100415

17. Auta A, Adewuyi EO, Kureh GT, Onoviran N, Adeloye D. Hepatitis B vaccination coverage among health-care workers in Africa: A systematic review and meta-analysis. Vaccine. 2018; 36:4851–60. 10.1016/j.vaccine.2018.06.043

18. Noubiap JJN, Nansseu JRN, Kengne KK, Tchokfe Ndoula S, Agyingi LA. Occupational exposure to blood, hepatitis B vaccine knowledge and uptake among medical students in Cameroon. BMC Med Educ. 2013; 13:148. 10.1186/1472-6920-13-148

19. Ngum AM, Laure SJ, Tchetnya X, Tambe TA, Ngwayu CN, Wirsiy FS, et al. Vaccination against Hepatitis B among health care workers in the Bamenda Health District: influence of knowledge and attitudes, Cameroon. Pan Afr Med J. 2021;40. 10.11604/pamj.2021.40.216.16856

20. Bilounga Ndongo C, Eteki L, Siedner M, Mbaye R, Chen J, Ntone R, et al. Prevalence and vaccination coverage of Hepatitis B among healthcare workers in Cameroon: A national seroprevalence survey. J Viral Hepat. 2018; 25:1582–7. 10.1111/jvh.12974

21. Cheuyem FZL, Mouangue C, Ajong BN, Edzamba MF, Hamadama DCM, Achangwa C, et al. Occupational exposure to blood and other body fluids among healthcare workers in Cameroon: A systematic review and meta-analysis. Glob Health Econ Sustain. AccScience Publishing; 2025; 3:185–96. 10.36922/GHES025090016

22. Takougang I, Cheuyem FZL, Lyonga EE, Ndungo JH, Mbopi-Keou F-X. Observance of Standard Precautions for Infection Prevention in The Covid-19 Era: A Cross-Sectional Study in Six District Hospitals in Yaounde, Cameroon. Am J Biomed Sci Res. biomedgrid; 2023; 19:590–8. 10.34297/AJBSR.2023.19.002628

23. Ogoina D, Pondei K, Adetunji B, Chima G, Isichei C, Gidado S. Prevalence of Hepatitis B Vaccination among Health Care Workers in Nigeria in 2011–12. Int J Occup Environ Med. 2014; 5:51–6.

24. Ngekeng S, Chichom-Mefire A, Nde PF, Tendongfor N, Nji EK, Malika E, et al. Hepatitis B Vaccination Coverage and Its Predictors among Health Workers in Fako Division, South West Region of Cameroon. Open Access Libr J. Scientific Research Publishing; 2022; 9:1–13. 10.4236/oalib.1108985

25. Orotta M, Munseri P, Massawe RV, Orotta GM, Ebrahim A, Kisali EP, et al. Hepatitis B virus: Prevalence, vaccination coverage and immune responses to immunization among healthcare workers at Muhimbili National Hospital. PLOS ONE. Public Library of Science; 2025;20: e0321623. 10.1371/journal.pone.0321623

26. Mueller A, Stoetter L, Kalluvya S, Stich A, Majinge C, Weissbrich B, et al. Prevalence of hepatitis B virus infection among health care workers in a tertiary hospital in Tanzania. BMC Infect Dis. 2015 ;15 :386. 10.1186/s12879-015-1129-z

27. Bigna JJ, Amougou MA, Asangbeh SL, Kenne AM, Noumegni SRN, Ngo-Malabo ET, et al. Seroprevalence of hepatitis B virus infection in Cameroon: a systematic review and meta-analysis. BMJ Open [Internet]. British Medical Journal Publishing Group; 2017 [cited 2026 Mar 27]; 10.1136/bmjopen-2016-015298

28. Bouba G, Fl A, Yj KF, E N. Prévalence de l’Hépatite B chez les Donneurs de Sang à Tokombéré (Extrême-Nord / Cameroun): Prevalence of Hepatitis B Among Blood Donors in Tokombere (Far North Region, Cameroon). Health Sci Dis. 2024; 25:92–8. 10.5281/hsd.v25i6.5772

29. Nzechieu Evenge CN, Zeuko’o Menkem E, Ngounou E, Watching D, Nembu EN, Luma WS, et al. Prevalence of hepatitis B and associated factors in the Buea Regional Hospital, Cameroon. Heliyon. 2023;9:e17745. 10.1016/j.heliyon.2023.e17745

