## Supplementary Material for "Vaccination status and associated factors among healthcare workers providing primary care: a cross-sectional study in Yaoundé, Cameroon"

**Date:** _____/____/2024	**Questionnaire N°**____________

*Instructions: Please circle the number corresponding to the correct answer.*

I. **SOCIO-DEMOGRAPHIC DATA**

1. **Age:** ______________________years

2. **Sex:** 1= Male 2 = Female

3. **Level of education:** 1 = primary 2 = secondary 3 = tertiary

4. **Grade:** 1 = Specialist doctor 2= General practitioner 3 = Nurse 4 = Midwife/midwife 5 =	Care assistant 6= Laboratory technician 7= Hygiene staff	8= other: ............

5. **Service where you work: ___________________________**

6. **Time already spent in this department**: ____________years _________ months

7. **Number of years of hospital service since training completion**: ________years ______ months

II. **VACCINATION COVERAGE AND COMPLIANCE**

**8**. **Have you ever been vaccinated against viral hepatitis B**? 1 = Yes 2 = No

**8.1 If yes, specify the number of doses received**: 1 = 1 dose 2 = 2 doses 3 = 3 doses

**8.2 If yes, date of last dose**: ................ (Months +/-Years)

**8.3 If yes, did you have any adverse reactions to the vaccination?** 1=Yes 2= No

**8.3.1 If yes, which ones?** 1= Fever 2= Headache 3= swelling of lymph nodes in armpit 4= large sore that took a long time to heal 5= other: please specify...............................................................................

**8.4 If you have not received this vaccine, what may be the cause** 1= Doubts about the real need for vaccines 2= Fear of side effects 3= Vaccine unavailable 4= High-cost 5= for religious or philosophical reasons 6= Negative experience with vaccines

**9.** **Have you ever been vaccinated against tuberculosis?** 1 = Yes 2 = No

**9.1 If yes, specify the number of doses received**: 1 = 1 dose 2 = 2 doses 3 = 3 doses

**9.2 If yes, date of last dose:** ................ (Months +/-Years)

**9.3 If yes, did you have any adverse reactions to the vaccination**? 1=Yes 2= No

**9.3.1 If yes,** which ones? 1= Fever 2= Headache 3= Swelling of lymph nodes in armpit 4= Large sore that took a long time to heal 5= other; please specify.................................................................................

**9.4 If you have not received this vaccine, what may be the cause?** 1= Doubts about the real need for vaccines 2= Fear of side effects 3= Vaccine unavailable 4= High cost 5= for religious or philosophical reasons 6= Negative experience with vaccines

**10**. **Have you ever been vaccinated against tetanus?** 1 = Yes 2 = No

**10.1 If yes, specify the number of doses received:** 1= 1 dose 2 = 2 doses 3 = 3 doses 4= 4doses 5=5doses

**10.2 If yes, date of last dose:** ....................................................... (Months +/-Years)

**10.3 If yes, did you have any adverse reactions to the vaccination?** 1=Yes 2= No

**10.3.1 If yes, which ones?** 1= Fever 2= Headache 3= Swelling of lymph nodes in armpit 4= Large sore that took a long time to heal 5= other, please specify................................................................................

**10.4 If you have not received this vaccine, what may be the cause**? 1= Doubts about the real need for vaccines 2= Fear of side effects 3= Vaccine unavailable 4= High cost 5= for religious or philosophical reasons 6= Negative experience with vaccines
